# Loss of smell and taste in combination with other symptoms is a strong predictor of COVID-19 infection

**DOI:** 10.1101/2020.04.05.20048421

**Authors:** Cristina Menni, Ana M Valdes, Maxim B Freidin, Sajaysurya Ganesh, Julia S El-Sayed Moustafa, Alessia Visconti, Pirro Hysi, Ruth C E Bowyer, Massimo Mangino, Mario Falchi, Jonathan Wolf, Claire J Steves, Tim D Spector

**Affiliations:** Department of Twin Research and Genetic Epidemiology, King’s College London, Westminster Bridge Road, SE17EH London, UK; Academic Rheumatology Clinical Sciences Building, Nottingham City Hospital, Hucknall Road, Nottingham, NG5 1PB, UK; Zoe Global Limited,164 Westminster Bridge Road, London SE1 7RW, UK

## Abstract

**Importance:** A strategy for preventing further spread of the ongoing COVID-19 epidemic is to detect infections and isolate infected individuals without the need of extensive bio-specimen testing.

**Objectives:** Here we investigate the prevalence of loss of smell and taste among COVID-19 diagnosed individuals and we identify the combination of symptoms, besides loss of smell and taste, most likely to correspond to a positive COVID-19 diagnosis in non-severe cases.

**Design:** Community survey.

**Setting and Participants:** Subscribers of RADAR COVID-19, an app that was launched for use among the UK general population asking about COVID-19 symptoms.

**Main Exposure:** Loss of smell and taste.

**Main Outcome Measures:** COVID-19.

**Results:** Between 24 and 29 March 2020, 1,573,103 individuals reported their symptoms via the app; 26% reported suffering from one or more symptoms of COVID-19. Of those, n=1702 reported having had a RT-PCR COVID-19 test and gave full report on symptoms including loss of smell and taste; 579 were positive and 1123 negative. In this subset, we find that loss of smell and taste were present in 59% of COVID-19 positive individuals compared to 18% of those negative to the test, yielding an odds ratio (OR) of COVID-19 diagnosis of OR[95%CI]=6.59[5.25; 8.27], P= 1.90×10^−59^. We also find that a combination of loss of smell and taste, fever, persistent cough, fatigue, diarrhoea, abdominal pain and loss of appetite is predictive of COVID-19 positive test with sensitivity 0.54[0.44; 0.63], specificity 0.86[0.80; 0.90], ROC-AUC 0.77[0.72; 0.82] in the test set, and cross-validation ROC-AUC 0.75[0.72; 0.77]. When applied to the 410,598 individuals reporting symptoms but not formally tested, our model predicted that 13.06%[12.97%;13.15] of these might have been already infected by the virus.

**Conclusions and Relevance:** Our study suggests that loss of taste and smell is a strong predictor of having been infected by the COVID-19 virus. Also, the combination of symptoms that could be used to identify and isolate individuals includes anosmia, fever, persistent cough, diarrhoea, fatigue, abdominal pain and loss of appetite. This is particularly relevant to healthcare and other key workers in constant contact with the public who have not yet been tested for COVID-19.

**Key points:** *What is already known on this topic:*

- The spread of COVID-19 can be reduced by identifying and isolating infected individuals but it is not possible to test everyone and priority has been given in most countries to individuals presenting symptoms of the disease.
- COVID-19 symptoms, such as fever, cough, aches, fatigue are common in many other viral infections
- There is therefore a need to identify symptom combinations that can rightly pinpoint to infected individuals

*What this study adds:*

- Among individuals showing symptoms severe enough to be given a COVID-19 RT-PCR test in the UK the prevalence of loss of smell (anosmia) was 3-fold higher (59%) in those positive to the test than among those negative to the test (18%).
- We developed a mathematical model combining symptoms to predict individuals likely to be COVID-19 positive and applied this to over 400,000 individuals in the general population presenting some of the COVID-19 symptoms.
- We find that ∼13% of those presenting symptoms are likely to have or have had a COVID-19 infection. The proportion was slightly higher in women than in men but is comparable in all age groups, and corresponds to 3.4% of those who filled the app report.

## Introduction

Since the outbreak of COVID-19, an acute respiratory illness caused by the novel coronavirus SARS-CoV-2, in mainland China in December 2019, there have been more than 700,000 confirmed cases worldwide ^1^. Global attention has been mainly on the infected patients and frontline responders, with little visibility of the hidden iceberg of cases of coronavirus that are not yet in hospital. Indeed, a large proportion of the UK (and world) population is presenting with flu like symptoms, but widespread population testing is not yet available in countries like the US^2^ and the UK^3^. It is therefore of upmost importance to identify the combination of symptoms that are most predictive of COVID-19 infection, in order to instruct on self-isolation and prevent the spreading of the disease.

Case and media reports from China, Italy, South Korea, France, Germany and most recently the UK ^4-6^ indicate that a significant number of patients with proven COVID-19 have developed anosmia (loss of smell) and a mechanism of action for the SARS-CoV-2 viral infection causing anosmia has been postulated elsewhere^7^. Moreover, there has been a growing number of reports indicating that a number of patients present anosmia also in the absence of other symptoms^8^. This is suggesting that anosmia could be used as a screening tool to help identify potential mild cases who could be instructed to self-isolate.

In the general population, 2-3% are affected by lifelong olfactory disorders (from hyposmia to anosmia)^9^ and two of the most frequent aetiologies are the common cold and flu^9^. Post-viral olfactory disorders usually occur after an upper respiratory tract infection (URTI) associated with a common cold or influenza^9^. Women are also more often affected than men and post-URTI disorders usually occur between the fourth and eighth decade of life^10^.

Here we investigate whether loss of taste and smell is specific to COVID-19 in 1,573,103 individuals who signed up to *COVID RADAR*, an app that tracks in real time how the disease progresses by recording health information on a daily basis, including temperature, tiredness and symptom. We first investigate the correlation between loss of smell and taste and COVID-19 in 579 cases and 1123 controls who had RT-PCR. We then identify which symptoms, besides anosmia, are most likely predicting COVID-19. Finally, we test our predictive model in 410,598 individuals who have symptoms but have not yet been tested.

## Methods

### Study setting and participants

The ***COVID RADAR*** Symptom Tracker app developed by Zoe Global Limited and King’s College London was launched in the UK on Tuesday the 24^th^ March 2020, and in less than a week has reached 1,573,103 subscribers. It enables capture of self-reported information related to COVID-19 infections. On first use, the app records self-reported location, age, and core health risk factors. With continued use, participants provide daily updates on symptoms, health care visits, COVID-19 testing results, and if they are self-quarantining or seeking health care, including the level of intervention and related outcomes. Individuals without apparent symptoms are also encouraged to use the app. Through direct updates, the research team can add or modify questions in real-time to capture new data to test emerging hypotheses about COVID-19 symptoms and treatments. Importantly, participants enrolled in ongoing epidemiologic studies, clinical cohorts, or clinical trials, can provide informed consent to link data collected through the app in a HIPPA and GDPR-compliant manner with extant study data they have previously provided or may provide in the future.

The research data will be shared with third parties submitting an application to the TwinsUK Research Executive Committee who will review and approve.

### Assessment of Exposure, Ascertainment of outcomes, Ascertainment of covariates

Exposure, outcome and covariates were all ascertained via the app. However, a subset of individuals was tested for COVID-19 with RT-PCR.

### Ethics

The App Ethics has been approved by KCL ethics Committee and all subscribers provided consent. An informal consultation with TwinsUK members over email and social media prior app been launched found that they were overwhelmingly supportive of the project.

### Statistical analysis

Data from the app were downloaded into a server and only records where the self-reported characteristics fell within the following ranges were utilised for further analyses: age between 16 and 90; height (cm) between 110 and 220; weight (kg) between 40 and 200; BMI(kg/m^2^) between 14 and 55; and temperature (in C) between 35 and 42.

Baseline characteristics are presented as the number (percentage) for categorical variables and the mean (standard deviation) for continuous variables.

Multivariate logistic regression adjusting for age, sex and BMI was applied to investigate the correlation between loss of taste and smell and COVID-19 in 579 cases and 1123 controls from participants of the RADAR COVID app who were also tested in the lab for COVID-19. In this same dataset, we then performed stepwise logistic regression combining forward and backward algorithms, to identify other symptoms associated to COVID-19 independently of loss of smell and taste. We included in the model ten other symptoms (including fever, persistent cough, fatigue, shortness of breath, diarrhoea, delirium, skipped meals, abdominal pain, chest pain and hoarse voice) as well as age and sex and chose, as the best model, the one with the lowest AIC. Prior to the modelling, to preserve the sample size, we imputed missing values for the symptoms of interest using missForest package in R ^11^]. The sample was randomly split into the train and test sets with 80/20 ratio. The model was fit using the train set and its performance was assessed using the test set. In addition, the performance of the model has been assessed via 10-fold cross-validation in the whole sample of 1703 individuals using the R package cvAUC^12^.

For our predictive model, using the R packages pROC and epiR, we further computed the area under curve (AUC) i.e. the overall diagnostic performance of the model, the sensitivity (“positivity in disease”) i.e. the proportion of subjects who have the target condition (reference standard positive) and give positive test results, and the specificity (“negativity in health”) i.e. the proportion of subjects without a COVID-19 RT-PCR test that give negative model results.

Finally, we applied the predictive model to the 410,598 individuals reporting symptoms who had not had a COVID-19 test in order to estimate the percentage of individuals reporting some COVID-19 symptoms likely to be infected by the virus. The proportion of estimated infections was calculated repeatedly by sampling the dataset (with replacement) to get the 95% confidence intervals.

## Results

The descriptive characteristics of all the “citizen scientists” are presented in **Table 1**. Briefly, 1,573,103 individuals filled the app report for symptoms between 24 and 29 March 2020, 26% of whom reported suffering from one or more symptoms of COVID-19. Out of these, 10.5% suffered from fever, 28.9% from persistent cough, 28.2% from shortness of breath, 53.1% from fatigue and 18.2% from loss of sense of smell or taste. We selected 1702 participants that reported (i) having had an RT-PCR COVID-19 test, (ii) having received the outcome of the test and (iii) symptoms including loss of smell and taste. Of these, 579 individuals tested positive for COVID-19 and 1123 tested negatively via RT-PCR.

**Table 1.**
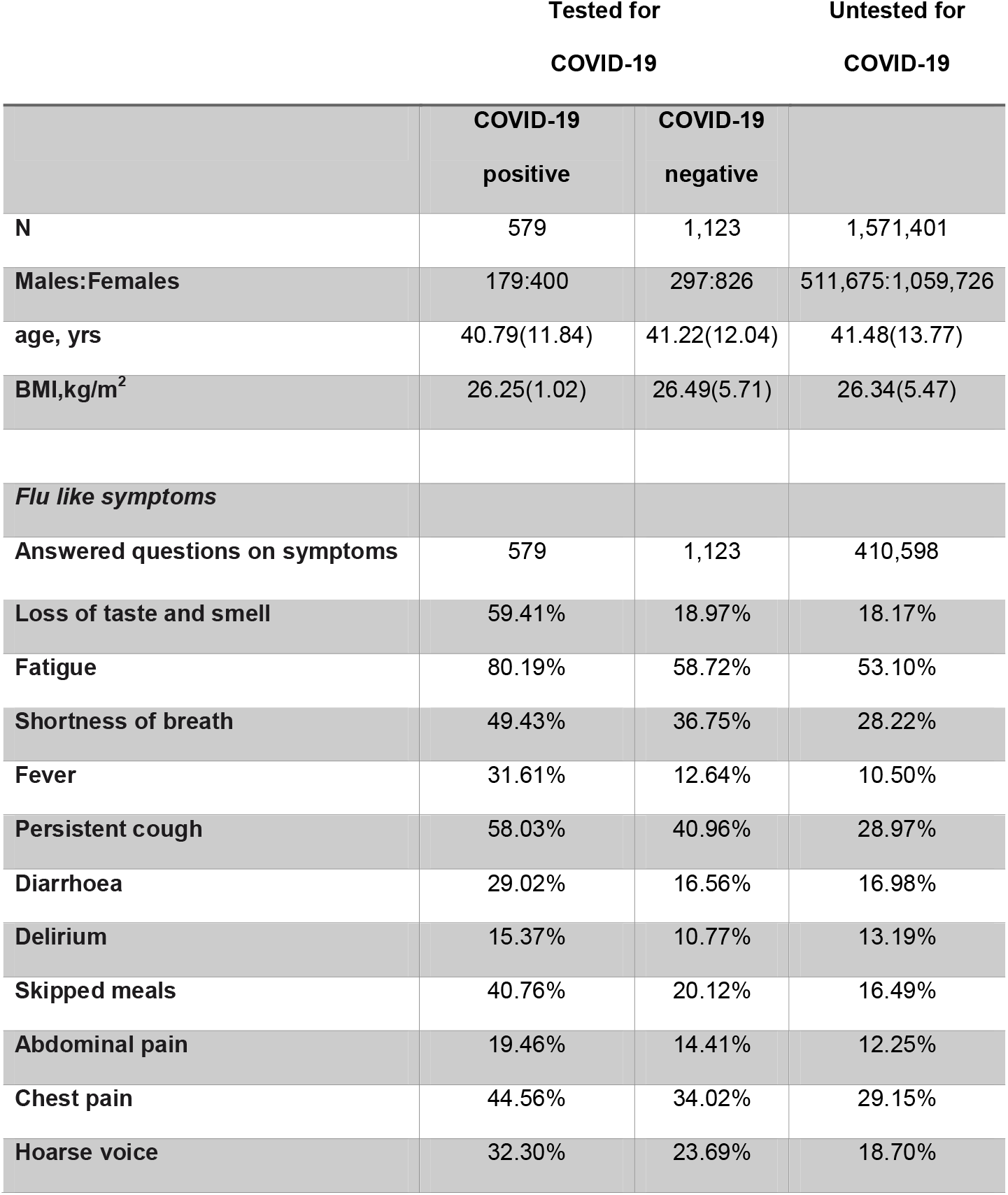
Descriptive characteristics of the study population. Results are presented as n(%) for dichotomous traits, as mean(SD) for continuous traits

In this subset, 59.41% of those testing positive for COVID-19 reported loss of taste and smell compared to only 18.97% of those who tested negative resulting in significantly higher odds of having being diagnosed with COVID-19 infection (OR[95%CI]= 6.59[5.25; 8.27], P= 1.90×10^−59^) after adjusting for age, sex and BMI. Overall, without any adjustments loss of sense of smell and taste has a positive predictive value of 61.7%.

We then reran a logistic regression adjusting for age, sex and BMI to identify other symptoms besides anosmia, associated with being infected by COVID-19. As shown in **Figure 1**, all the ten symptoms (fever, persistent cough, fatigue, shortness of breath, diarrhoea, delirium, skipped meals, abdominal pain, chest pain and hoarse voice) were positively associated to an increase odds of having COVID-19 after adjusting for multiple testing using False Discovery Rate.

**Figure 1.**
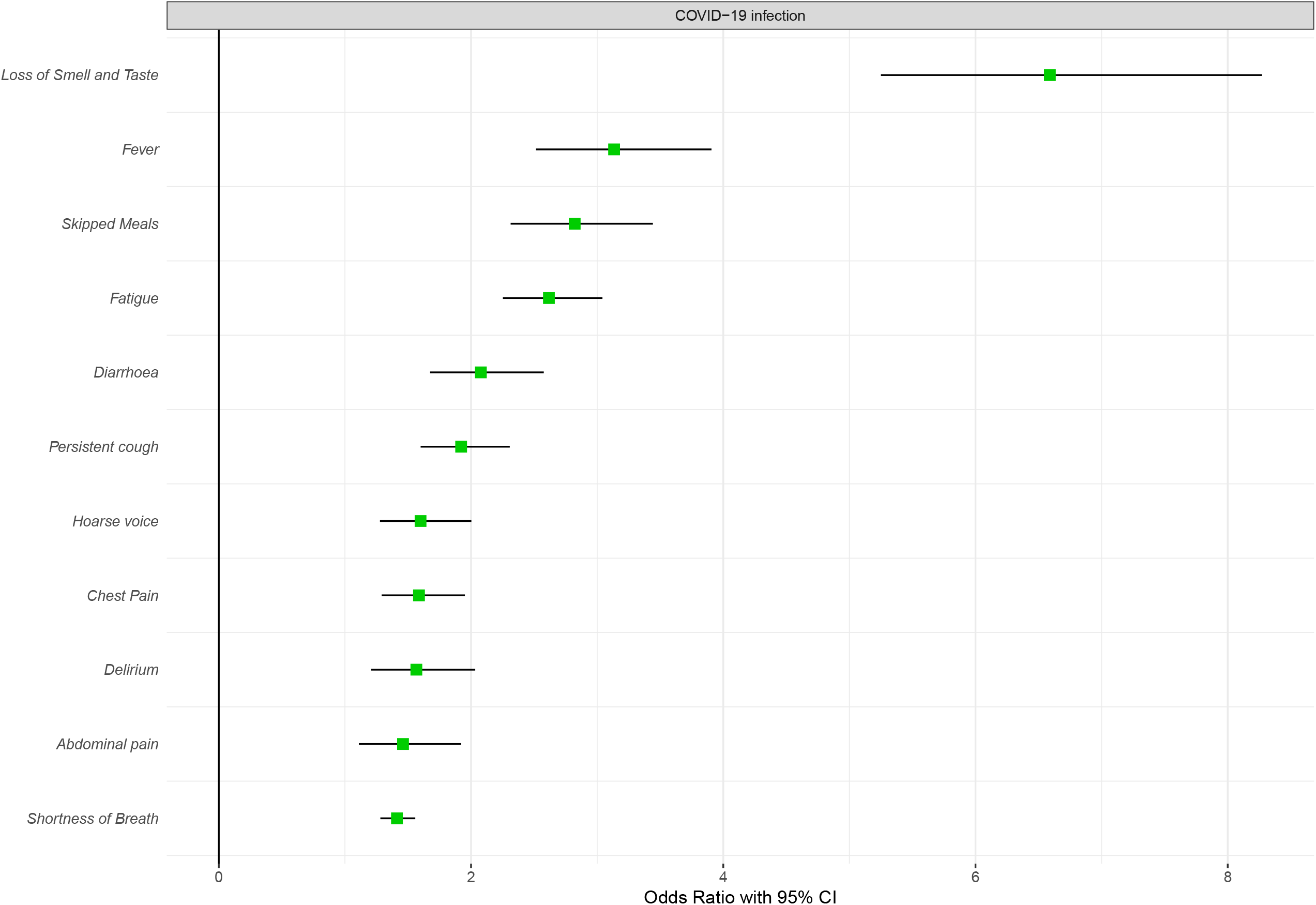
Association between symptoms and COVID-19 infection in 1702 participants that were tested via RT-PCR, OR(95%CI) are reported.

The Spearman correlation between loss of smell and taste and the other symptoms is presented in **Figure 2**, in the overall cohort, stratifying by sex, by age group, by COVID-19 test and diagnosis.

**Figure 2.**
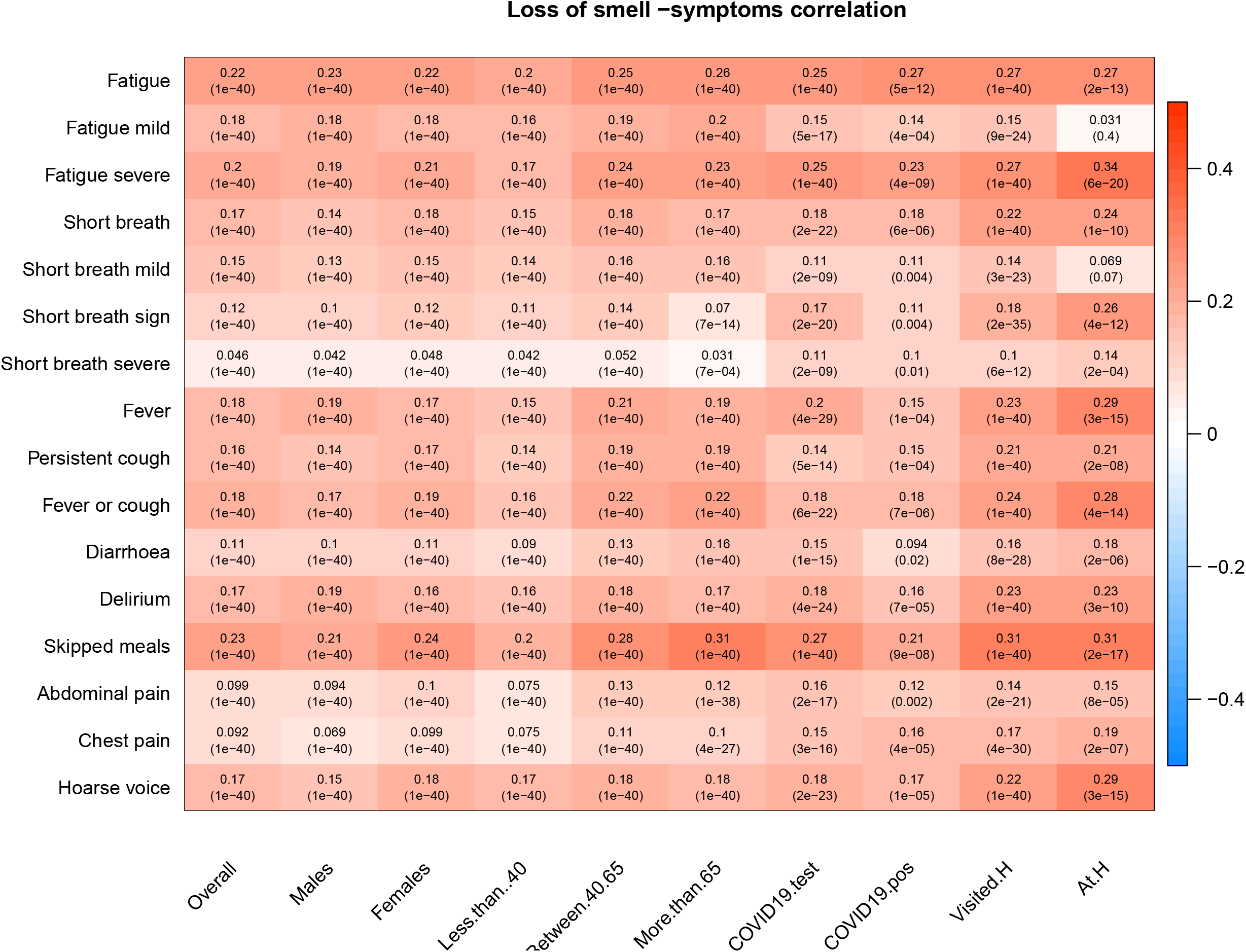
Spearman correlations between loss of smell and taste versus other flu symptoms in the subscriber of the COVID RADAR app and stratifying by different group of people. Each cell of the matrix contains the Spearman correlation coefficient between loss of taste and smell and one of the symptoms and the corresponding P value. The table is colour coded by correlation according to the table legend (red for positive and blue for negative correlations).

We then performed stepwise logistic regression to identify independent symptoms most strongly correlated to COVID-19 adjusting for age, sex and BMI. A combination of loss of smell and taste, fever, fatigue, persistent cough, diarrhoea, abdominal pain and loss of appetite resulted in the best model (with the lowest AIC). We therefore generated a linear model for symptoms that included loss of smell and taste, fever, fatigue, persistent cough, diarrhoea, abdominal pain and loss of appetite to get a symptoms prediction model for COVID-19:

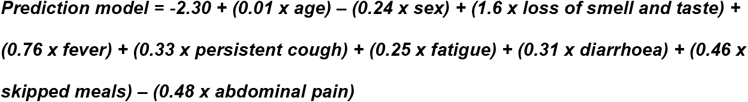

where all symptoms should be coded as 1 if the person reports to suffer from it and 0 if the person reports not suffering from it. The ‘sex’ feature is also binary, with 1 representing males and 0 representing females.

The prediction model had a sensitivity of 0.54[0.44; 0.63] and a specificity of 0.86[0.80; 0.90], and a ROC-AUC 0.77[0.72; 0.82]. Cross-validation ROC-AUC was 0.75[0.72; 0.77].

In this model, the strongest predictor was loss of smell and taste, **Figure 3a**. We also computed the ROC-AUC stratifying for sex (**Figure 3 panel b and c**) and age-groups (**Figure 3, panels d, e and f**) and found that results were similar in all groups with no significant differences between strata suggesting that our model works in the same way within different sex and age-groups.

**Figure 3.**
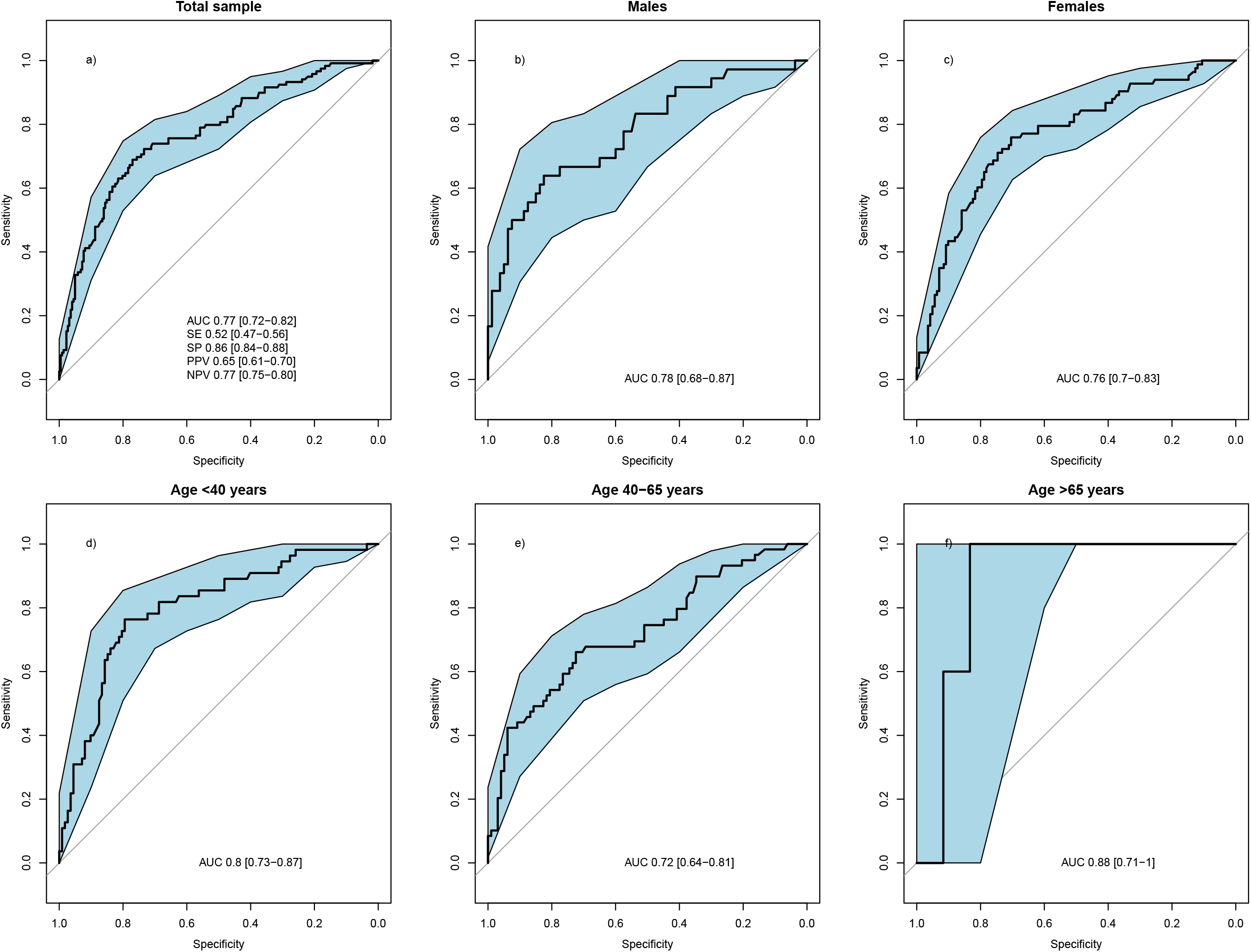
ROC-curve for prediction in the test set of the risk of positive test for COVID-19 using self-reported symptoms and traits: fever, persistent cough, fatigue, diarrhoea, skipped meals, abdominal pain, loss of smell, sex, age, and BMI in the overall COVID-19 tested subset and stratifying by sex and age groups.

Finally, we applied the predictive model to the 410,598 individuals reporting symptoms who had not had a COVID-19 test and we find that according to our model 13.06%[12.97%;13.15%] of individuals reporting some COVID-19 symptoms are likely to be infected by the virus. We have repeated the analysis stratifying by sex and age groups (see **Figure 4**). This adds up to 53,609 individuals, which represents 3.4% as proportion of the overall responders to our app.

**Figure 4.**
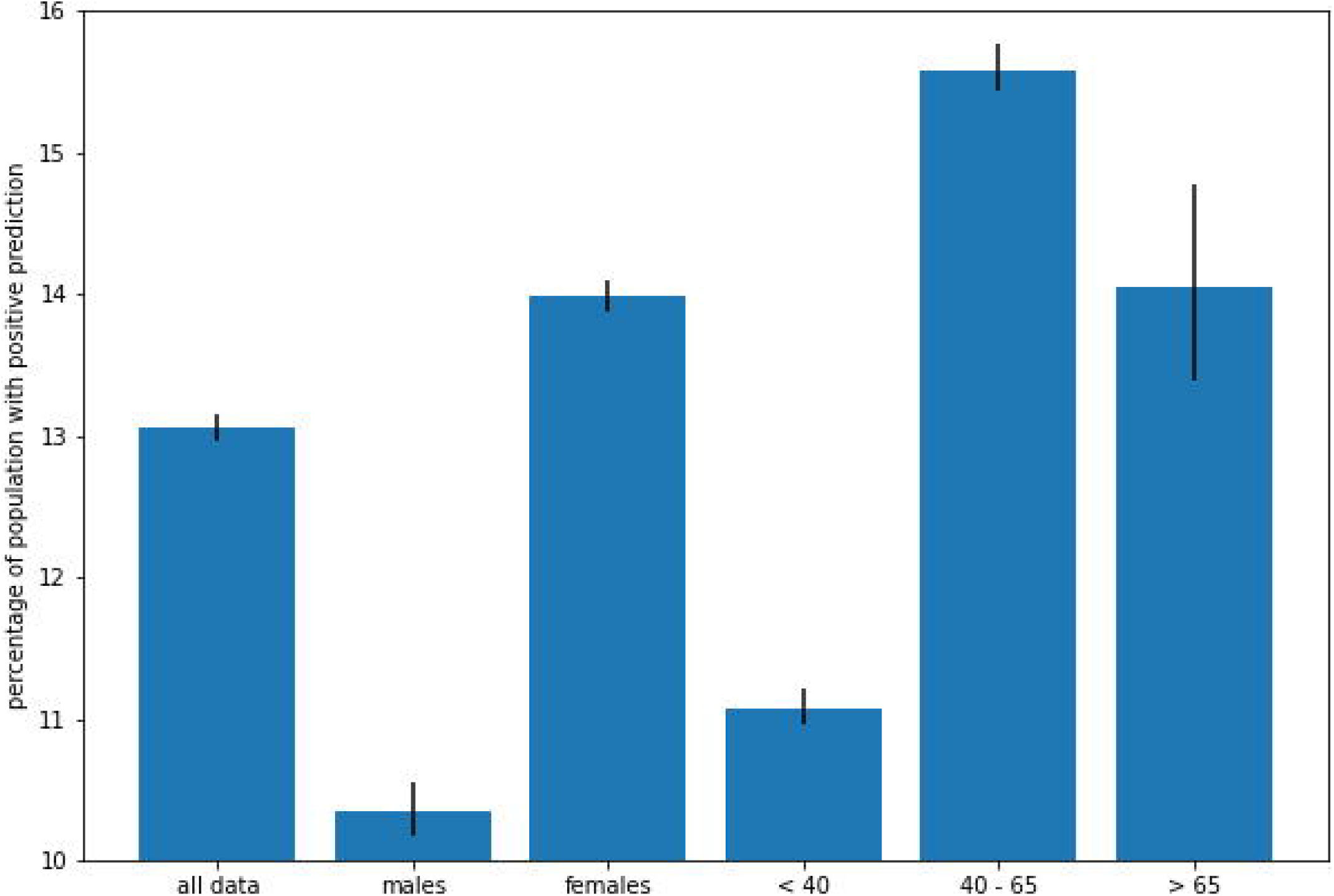
Proportion of participants untested for COVID-19 with an estimated probability of infection greater than 0.5. The proportion of estimated infections was calculated repeatedly by sampling the dataset (with replacement) to get the 95% confidence intervals. The analysis was repeated stratifying by sex and age groups.

## Discussion

Here we report that loss of smell and taste is a strong predictor of COVID-19 besides the most established symptoms of having a high temperature and a new, continuous cough. COVID-19 seems to cause problems of smell receptors in line with many other respiratory viruses, including previous coronaviruses thought to account for 10-15% of cases of anosmia^8^.

We also identify a combination of symptoms including anosmia, fever, fatigue, persistent cough, diarrhoea, abdominal pain and loss of appetite that together can identify with a high specificity and average sensitivity COVID-19 infected individuals.

Our study also has some limitations. First, all the data collected by us is self-reported. Second, at the moment, we don’t know whether anosmia was acquired prior to other COVID-19 symptoms, during the illness or afterwards. This information will become available as the App actually tracks over time. Thirdly, the COVID-19 diagnosis is based on the RT-PCR test that has less than 100% sensitivity (true positive rate)^13^. More accurate tests would provide a more accurate diagnosis and our results might be slightly different. An important caveat is that the individuals on which the model was trained are highly selected because RT-PCR COVID-19 tests are not random. It is that they were tested because they either displayed severe symptoms, were in contact with Covid-19 individuals health workers, or travelled in an area of particular risk. Therefore, we may be overestimating the number of expected positives. Also, the estimates on the prevalence of COVID-19 derived from our model and from self-reported symptoms derived from the members of the public who took part in the app. This has self-selected a group not fully representative of the general population as the sex proportions of our respondents clearly show, with women being much more likely to respond than men and people under 60 representing the majority (>80%) of responders.

Our data suggest that anosmia should be added by the World Health Organisation to their COVID-19 symptom list^14^. In the presence of other symptoms, people with loss of smell and taste appear to be 3 times more likely to have contracted the virus according to our data and should therefore self-isolate for fourteen days to reduce the number of people who continue to act as vectors.

The current research suggests that loss of sense of smell and taste could be implemented as part of the screening for COVID-19, however its usefulness in a public health setting will depend on whether this symptom occurs early in the onset of symptoms or only after more severe symptoms, such as cough, high fever and shortness of breath are present. A detailed study on the natural history of COVID-19 symptoms, the order and frequency with which they occur will help answer these questions. A second question is for how long after a person has cleared the virus does this symptom persist and whether it is mostly temporary as is the case for anosmia from other viral infections, or if it has a longer persistence in time.

## Acknowledgments

We express our sincere thanks to the participants of the RADAR COVID app. We thank the staff of Zoe Global Limited, the Department of Twin Research for their tireless work in contributing to the running of the study and data collection.

## Authors contribution

Dr Menni and Prof Spector had full access to all of the data in the study and take responsibility for the integrity of the data and the accuracy of the data analysis. Drs Menni and Valdes contributed equally to this manuscript. Dr Steves and Prof Spector also contributed equally to the study.

***Conceived and designed the experiments*:** CJS, TDS; ***Analyzed the data*:** CM, MBF, SG, AMV. ***Contributed reagents/materials/analysis tools***: MBF, SG, AV, JESM, PH, MM, MF. ***Wrote the manuscript***: CM, AMV. ***Revised the manuscript***: all

The corresponding authors attests that all listed authors meet authorship criteria and that no others meeting the criteria have been omitted.

